# Development of a Community Orientation Program (COP) as a Community-Based Medical Education method for undergraduate medical students: An experience from India

**DOI:** 10.1101/2021.02.07.21251321

**Authors:** Bayapa Reddy Narapureddy, Shakeer Patan Kahn, C Sravana Deepthi, Sirshendu Chaudhuri, KR John, Chittooru Chandrasekar, Surendra Babu, Khadervali Nagoor, Devika Jeeragyal, Basha K. Jawahar, Theo Nell

## Abstract

**Background:** Intraregional cultural and linguistic differences are common in the Low- and Middle-income countries. They must focus on innovative teaching for the undergraduate (UG) medical students to make them sensitive to the social and contextual determinants of health to achieve the health for all goal. The early introduction of community-based medical education could be a game changing strategy.

**Objectives:** To describe the methods, evaluation, implication, and challenges of a Community Orientation Programme (COP) adopted for the Indian Medical Graduates (IMG). Methods: The COP was arranged for the first-year UG students, including the community and the local administration. The program was divided into phases like-initial preparation, theoretical sessions, field visits, group activities, data analysis, and dissemination of the findings. In this learner-centric, supervised educational program, the key aim was to focus on developing students’ communication skills, observation power and enhancing their motivation for learning new things through collaborative learning. A mixture of adult learning theories like behaviorism, cognitivism, constructivism, sociocultural theory, critical theory, and humanism included keeping the students’ needs, cultural differences, and varying motivational levels. A structured feedback mechanism from the students was developed to identify the gaps.

**Conclusion:** The COP has provided a holistic learning framework for the students based on several complementary learning theories in the Indian context. It has touched upon most of the national- and institutional-level objectives envisioned by the regulatory body for the IMGs. All stakeholders should promote such programs and solve the challenges collectively.

## Introduction

Community-based medical education (CBME) is an integral part of the current undergraduate (UG) medical curriculum. It complements classroom and clinical teaching by introducing learning within the community. (1–3) Early introduction of CBME in UG medical education helps the students to understand the social determinants of health and the contextual factors that are associated with various health conditions. It also enhances the socio-humanistic skills like communication, collaboration, (Dolmans, Dornan) listening, observation, leadership, and clinical skills, and supports the development of decision making abilities through group activities. (4,5) With CBME, the students get the opportunity to be exposed to the community they are going to serve in the future, develop the rapport with the community, work as a team, and get the opportunity to learn about various local prevailing diseases. (6,7) CBME has an inherent ability to identify and adapt with the changing characteristics of the community, like-socio-political situation, environmental condition, and disease trend to remain as a pillar of undergraduate medical teaching. (8,9)

Undergraduate medical teaching in India is often criticised for their knowledge-based learning rather than focussing on competency-based learning. (10–13) However, the Medical Council of India (MCI), the apex regulatory body for medical teaching in India, has restructured the undergraduate curriculum recently with an aim to develop core competencies amongst students. (14) The MCI has envisioned that every student should be a clinician, life-long learner, communicator, leader, and should act professional in his/her academic and professional life. (14,15) The MCI has also emphasized developing key sub-competencies like attitude, ethics, and communication skills amongst the Indian Medical Graduates (IMG) from commencing their undergraduate studies. (14,15) To meet this changing need in medical teaching, more innovative ideas in medical teaching should be fostered. Although CBME is commonly practiced (8), the experiences of these programs are not adequately shared in scientific literature.

The Community Medicine Department of a private medical college in South India started a CBME program, named the Community Orientation Program (COP) for the first semester students from 2016, which is also the inception year of the college. Since 2018, the COP became a part of the foundation course as laid down by the Graduate Medical Regulations laid down by the MCI. The college is situated in a semi urban part of Andhra Pradesh state of India. The college is supported by the government district hospital for clinical teaching. The college has a capacity of 150 undergraduate (UG) medical students per academic year. The Community Medicine Department runs various clinical and academic activities through initiatives like a mobile health clinic in the hard to reach areas, school health program, and field-based training of students through Urban and Rural Health Training Centres. It also conducts various field-research activities. To date the college have conducted four COPs up until the end of 2019. In this paper we describe the methods, evaluation, and implication of the COP.

## Method

The Institutional Curriculum Committee allocated 60 hours of training −30 hours theoretical and 30 hours practical training -for the Community Medicine Department, as prescribed by the Medical Council of India. With inputs from both teaching and non-teaching staff of the Department, a ‘Community Orientation Program’ (COP), was introduced (Box1) to complement the classroom teachings.

Every year, the COP is completed in four stages -planning, preparation, implementation, and feedback (Box-2).

### Box 1: Objectives of the COP

At the end of the COP, students are expected to:

1. Recognize the community structure
2. Observe the various cultures prevailing in the community
3. Develop communication skills
4. Perform basic clinical examinations
5. Recognize the various determinants of health, for example social-economic status, environmental condition, and social factors
6. Identify various diseases and clinical conditions prevailing in the community
7. Describe the needs of the community

### Box 2: Stages of the COP

I. **Planning stage:**
  A. Developing the concept proposal
  B. Involving the stakeholders and finalizing the plan
  C. Approval for the COP
II. **Preparatory stage**
  A. Administrative preparation
    - Selection of areas
    - Permission from local authority
    - Transport
    - Instruments
    - Food/ Refreshments
  B. Academic preparation
    - Social mapping
    - Development of proforma
    - Development of proforma filling guidelines
    - Training of the faculty and team
    - Division of roles and responsibilities
III. **Implementation stage**
  A. Student briefing, which is classroom-based
  B. Grouping & assignment
  C. Field activities
  D. Data entry and analysis
  E. Presenting the findings
IV. **Feedback**

### Planning stage

During the Departmental discussions, two major factors which influenced the program were kept in mind, namely that the students are new to the field of medicine, and that they have limited medical knowledge. Based on these factors, as well as considering aspects like the different motivational level of the students at the beginning of the program and an unknown teaching environment for them, a decision was taken to provide them with only the basic theoretical knowledge before introducing them to the community. (Figure 1) A key aim was to focus on the students’ communication skills, observation power, and enhancing their motivation for learning new things through collaborative learning. (Box-1)

**Figure 1:**
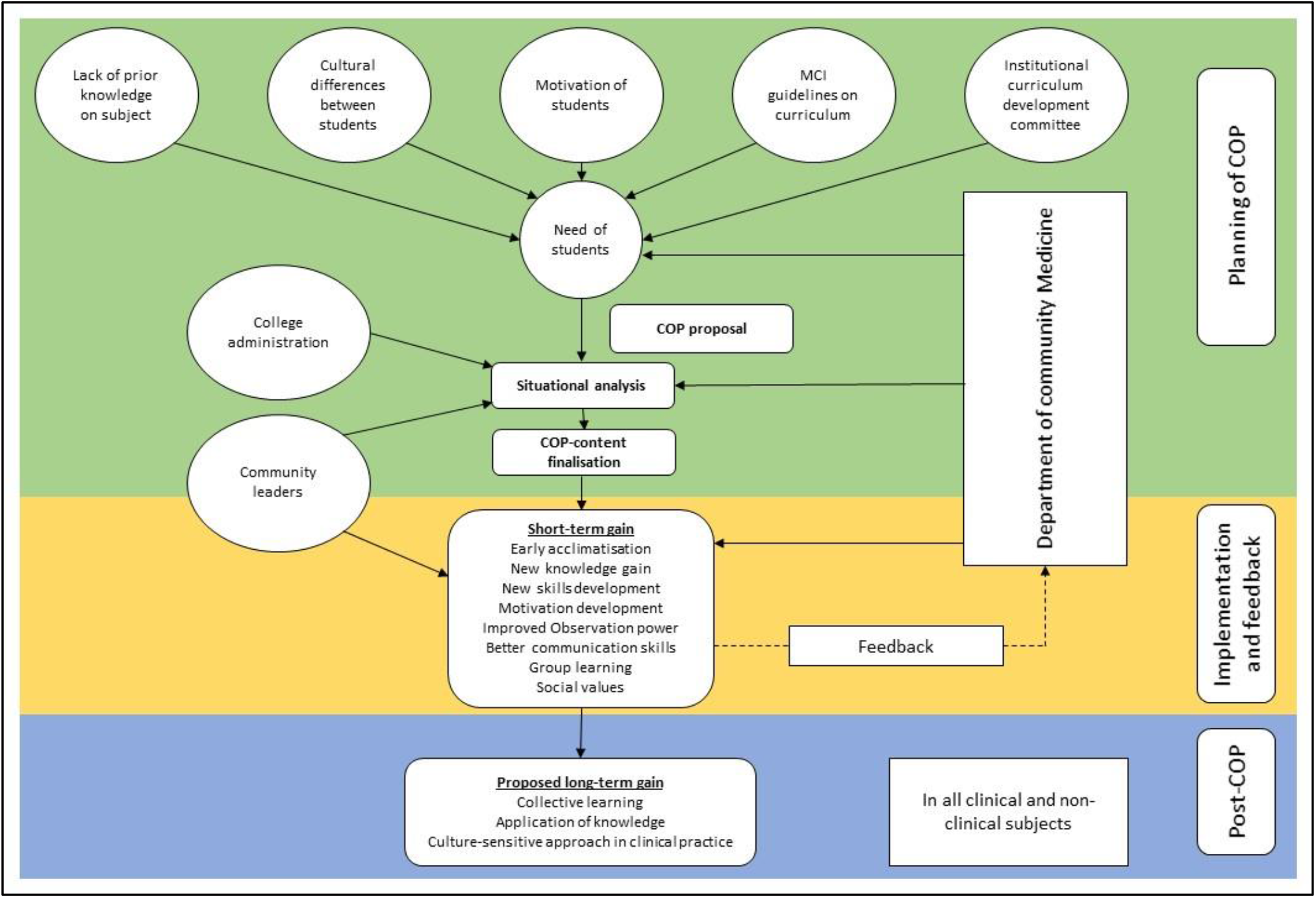
Framework of the COP

To deliver on all the objectives, the advantages, and disadvantages of several adult learning theories were discussed and included behaviourism, cognitivism, constructivism, sociocultural theory, critical theory, and humanism. (16–19) Considering the wide scope of the COP, a mix of various approaches were used rather than using only one approach. Based on the needs of the students, their cultural differences and varying motivational level, a micro-curriculum were followed which allowed for structured feedback from the students which influenced the possibility to restructuring and adapt the program. (Figure 1) After multiple discussions with the Institutional Curriculum Committee of the college, the college administration, as well as community leaders, the Institutional Head approved the content of the COP. (Table 1)

**Table 1:** Content of COP

| Topic covered | Areas covered | Duration (Hours) | Domain | Level of competency (Miller's pyramid) | Teaching learning method | Learning theory applied |
| --- | --- | --- | --- | --- | --- | --- |
| Concept on Demography (T) | Basic demographic and fertility indicators | 3 | Knowledge | Knows | Lecture/ interactive method | Cognitivism |
| Concept of social science and health (T) | Social and cultural factors in health and disease, Structure, and role of family in health in Indian context, socio-economic status classification systems, behavioural psychology | 5 | Knowledge | Knows | Lecture/ interactive method | Cognitivism |
| Environment and health (T) | Housing standards, waste disposal in community, biomedical waste management | 3 | Knowledge | Knows | Lecture/ interactive method | Cognitivism |
| National health programs related to common communicable and non-communicable diseases (T) | National Health Policy, National Population Policy, Universal Health Coverage | 3 | Knowledge | Knows | Lecture/ interactive method | Cognitivism |
| Indian Health System (T) | Structure and function of Indian Health system | 1 | Knowledge | Knows | Lecture/ interactive method | Cognitivism |
| Demonstration of proforma (T) | How to fill the data collection proforma | 4 | Knowledge | Knows/ Knows how | Demonstration/ interactive method | Behaviourism/ Cognitivism |
| Clinical examination (T and P) | Basic clinical examinations | 2 | Skill | Knows/ Knows how/ Skill/ Shows how | Demonstration |  |
| Interview and clinical exam (P) | Interviewing members of allocated families | 9 | Attitude, skill, communication | Does | Observation | Socio-cultural |
| Group discussion (T and P) | On various observations and clinical findings | 6 | Knowledge, attitude, communication | Knows/ Knows how | Small group teaching/ Peer teaching/ Collaborative teaching | Constructivism |
| Preparation of health education materials and health education session (T and P) | Diet, lifestyle modifications, and treatment compliance in NCDs | 3 | Knowledge, Skill, Communication | Knows/ Knows how/ Skill/ Perform independently | Peer teaching/ Collaborative teaching/ interactive method | Constructivism/ Socio-cultural |
| Data entry/ analysis (P) | - | 4 | Knowledge, Skill | Knows/ Knows how | Demonstration | Cognitivism/ Constructivism |
| Presentation (P) | Theme based | 6 | Communication/ Skill | Knows/ Knows how/ Skill | Interactive method | Critical theory |

T-Theory, P-Practical

#### Box 3: Logic model of the COP

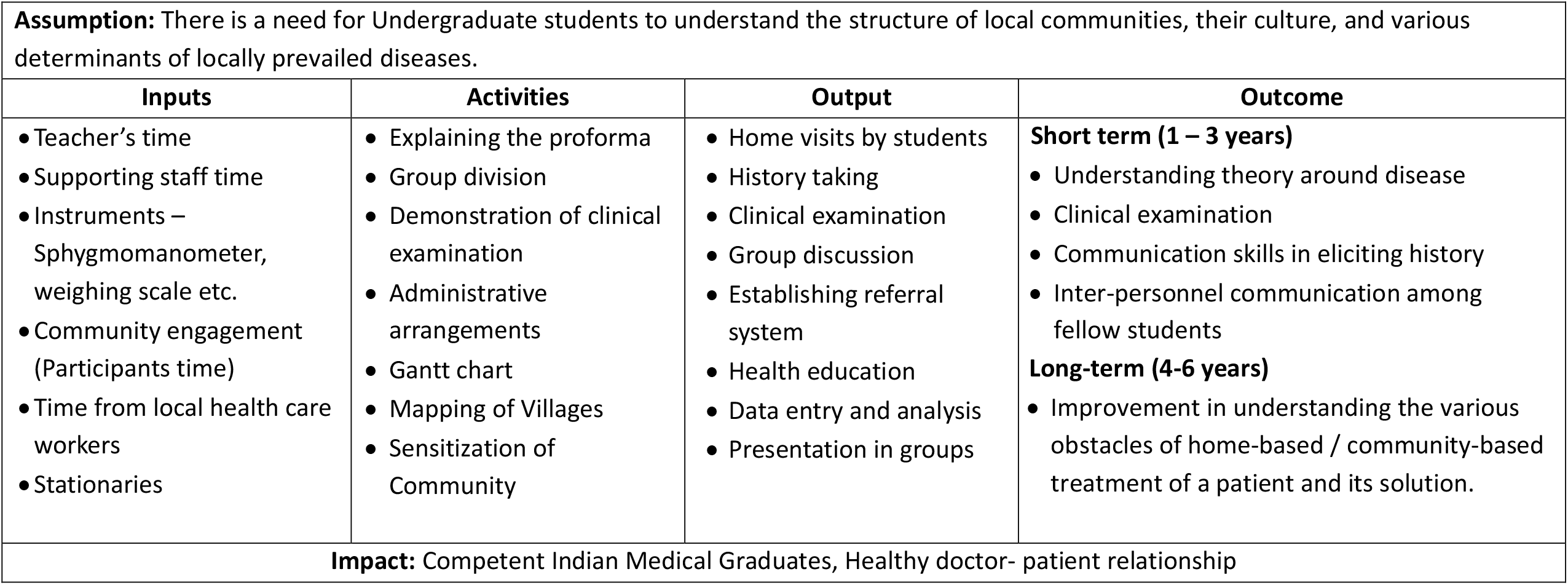

### Preparatory stage

The preparatory stage was divided into two parallel components, namely 1) the administrative preparation and 2) the academic preparation. A logic model, which summarised the inputs, activities, outputs, and projected outcomes, was prepared (Box 3) to enable a clear understanding on the various indicators of the COP.

#### Administrative Preparation

Administrative preparation includes-selection of village where the COP will be conducted, obtaining permission from the local authorities, and seeking local support for various administrative and academic support.

The tools and instruments needed for the program were purchased through funding provided and sanctioned by the Institute. In addition to the teaching and administrative staff, other departments like Transport, Food, and Information Technology (IT) were included to ensure the smooth coordination of the program.

#### Academic Preparation

Academic preparation includes-social mapping by the social workers (Figure 2) with the help of key informants in the village, and preparation of the interview proforma in English and in local vernacular with instructions for completing the proforma. The proforma is divided into various sections, and includes demographic characteristics, socio-economic conditions, environmental conditions (including housing conditions, water, kitchen, and sanitation), individual health condition (including-presence of any known disease(s), disability, pregnancy status (if applicable)), and clinical examination of the family members. From 2018, the paper format was replaced by an android-based electronic form. Each year, a nominated senior faculty member divides the roles and responsibilities among the other teaching and non-teaching staff.

**Figure 2:**
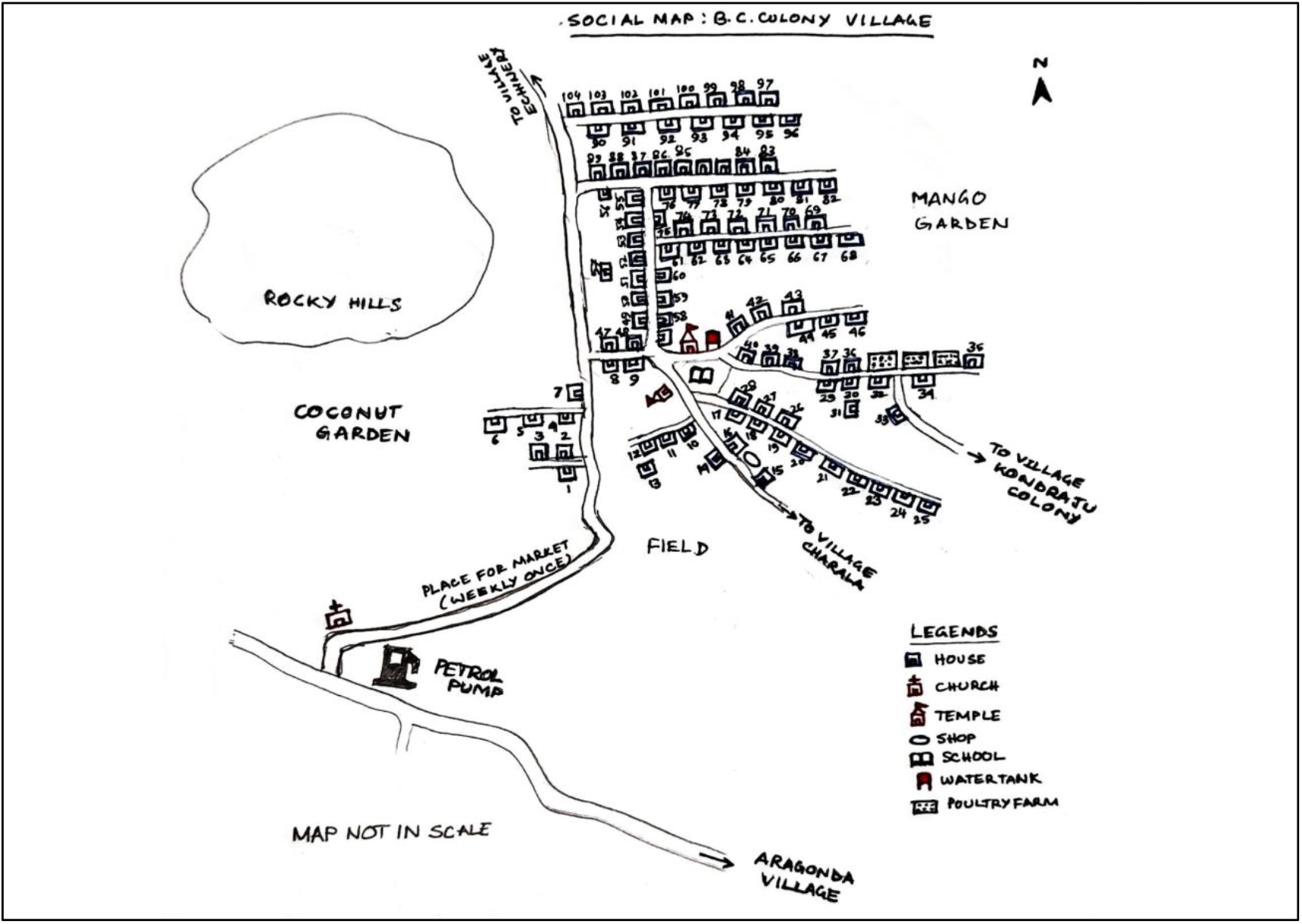
Social map of a selected village for COP

### Implementation phase

The implementation phase was divided into two distinct phases, namely 1) the theory sessions, and 2) the field visits (Figure 3)

**Figure 3:**
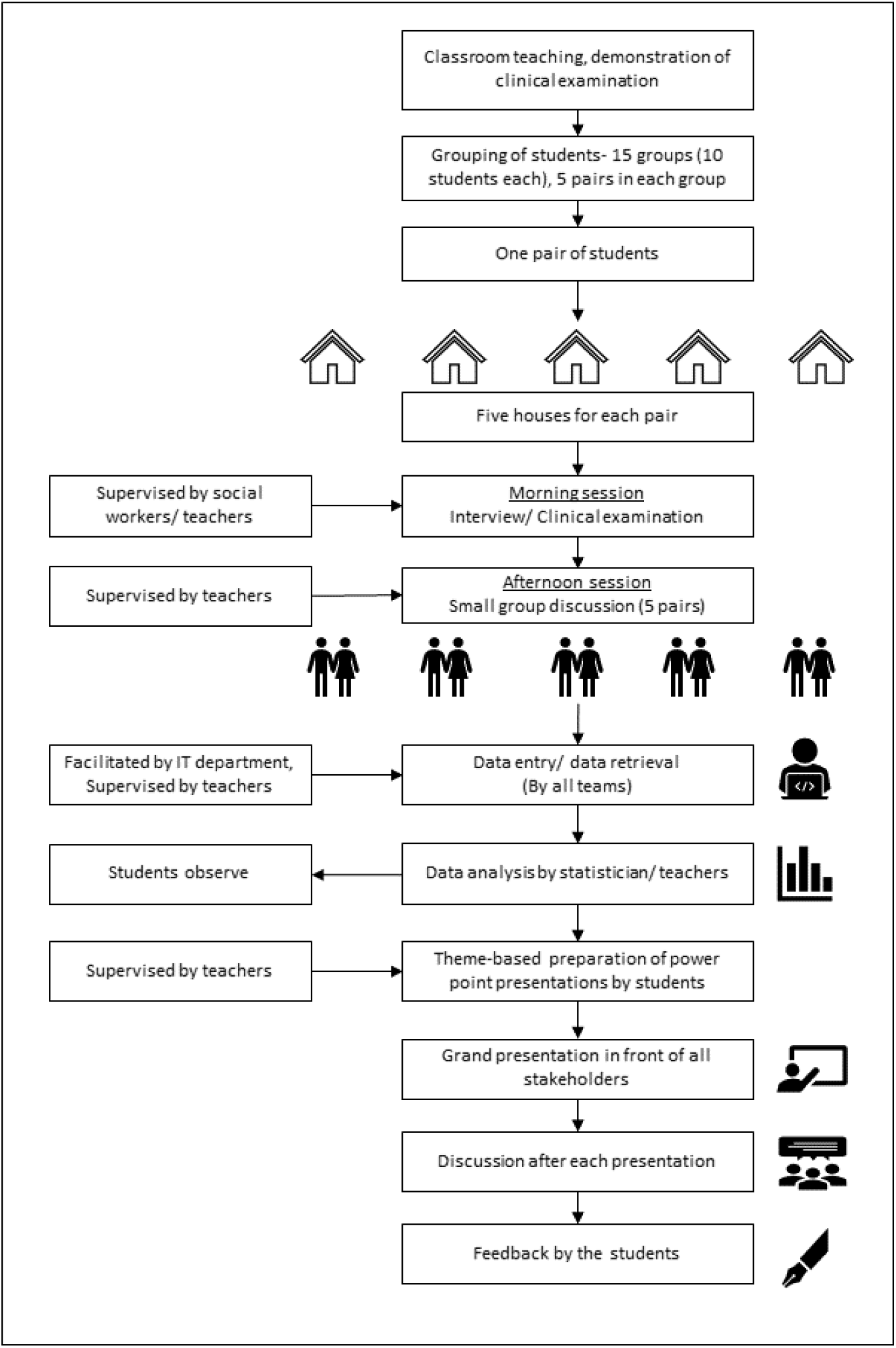
Implementation of the COP

#### Theory sessions

These included lectures with interactive sessions facilitated by two to three teachers in each session. In few sessions like teaching clinical examinations, theoretical knowledge and demonstrations were done simultaneously. (Table 1) An explanation of the field visit process, which included the proforma and how to complete it, was then done. Students were encouraged to conduct one mock interview with one of their fellow students before visiting the villages. This ensured that they were familiar with the proforma.

#### Field visits

Students were divided into fifteen groups of ten students per group, with each group under the supervision of one teacher. Within each group, students were divided into five pairs. Each pair was allocated five houses where interviews had to be conducted. (Figure 3) Social workers supported in the allocation of the houses, based on the line list done during the social mapping. In most of the cases pairs were formed to include at least one student who is well versed in the local vernacular, has an android device to use the online electronic form, and is female. This was done to ensure a balance in collecting sensitive information from the interviewees. If a pair did not have an android device, paper forms were given to them.

Each pair conducted the interview sessions with verbal consent of an adult interviewee of each allocated house, performed the clinical examination, and provided health education in relation to a balanced diet, the need of physical exercise, and importance of regular medical care if someone was already diagnosed with lifestyle disease(s). The social workers and the teachers directly supervised the interview sessions. If required, the teachers ensured an extensive clinical examination and provided advice on medicines or referred the interviewee to a government hospital.

During the afternoon small-group sessions, each group of five pairs interacted with each other under the guidance of the teacher. The various observations made in the community were discussed and questions were dealt with by the teacher. The teacher also explained the relationship of various socio-demographic and cultural habits with the diseases they came across during interview sessions. Students were also allowed to share their own experiences to help answer questions from their fellow students. On the final day of the field visit, a selected number of students performed health education activities, which included a role play and skit at a common place in the village to reinforce the health education provided at the family-level during the interview sessions.

#### Data entry and analysis

The computers in the central library of the institute were utilized with the support of the IT Department and the library staff. During 2016 and 2017, students entered data into excel spreadsheets under the supervision of a statistician and other teachers. After the introduction of the electronic questionnaire, the data was retrieved directly from the web. Based on fifteen different themes, the data was then analysed by the teachers while each group was allocated one theme. The students observed the analysis. The teachers also explained the fundamentals of basic analyses and how to calculate and interpret the data.

#### Presentation of findings

Each group presented their findings under the theme allocated to them to the teaching and administrative staff by using a PowerPoint presentation. Some also used audio-visual modes to explain their observations. The Department took utmost care to ensure anonymity of the respondents during the presentations. After the presentation of each theme, the presenting group, with the aid of their teacher, clarified questions posted by other groups, and teachers. The COP in charge summarized the findings at the end of the session. All students were expected to submit their collated findings to the Department.

### Evaluation of the COP

From 2016 to 2019, a total of 557 students enrolled in the medical program, of which 320 (57.5%) were female. All students participated in the COP. The input- and output-related indicators are shown in Table-2. In 2016, the inception year, the input-related lacunae were prominent. After completing the first COP, the team prepared the list of items required and human resource needs which were partially fulfilled in the subsequent years by the college administration, based on the available funding.

During the four-year period, a total of 4910 people from 1370 households in ten villages were surveyed. Out of the total population, 2576 (52.5%) were female, 263 (5.4%) were under five-year old children, and 559 (11.4%) were elderly people (>60 years).

Four hundred and one had chronic diseases, and fifty were pregnant women. (Table 2) The notable chronic diseases included diabetes mellitus, hypertension, physical disabilities (blindness, paralytic conditions due to poliomyelitis, Hansen’s disease, cerebrovascular accidents, and road traffic injuries), mental illnesses, and cancers of various origin. Among the acute cases, acute diarrhoeal and respiratory illnesses among under-five year old children and acute exacerbation of bronchial asthma were identified by the students. The acute conditions were either treated or referred after examination by the teachers, and students only played an observer role.

**Table 2:** Input and outcome evaluation of COP

| Resources utilised | Number/ duration |
| --- | --- |
| *Inputs related* |  |
| Teaching staff | Fifteen teachers (Three senior teachers, six junior teachers, four junior resident doctors, two medical officers including one gynaecologist, one statistician). In 2016, there were three junior teachers available. |
| Non-teaching personnel | • Three medical social workers, and one lab technician (Intradepartmental)<br>• Bus drivers: Four in 2016, five from 2017 onwards<br>• Village volunteers: three to five in each COP<br>• Village health workers: three to five in each COP |
| Instruments | • Weighing scale: five in 2016, 15 from 2017 onwards<br>• Sphygmomanometer- five in 2016, 15 from 2017<br>• Stethoscope: One for each student<br>• Measuring tape: One for each student |
| Vehicles | Buses: four in 2016, five from 2017 |
| ***Output-related*** |  |
| Implementation of COP | • Number of villages covered: 10<br>• Number of households covered: 1370<br>• Number of people covered: 4923<br>• Number of group discussion in the afternoon sessions: 157<br>• Number of common health education programs conducted: 10<br>• Number of groups presented in the final presentation: 57 |

#### Feedback from the students (Figure 4)

A survey on the students (n=332, response rate 60%) revealed that there are few areas where more improvement is needed. These include-demonstration of the clinical examinations (question 2), availability of adequate instruments for basic examinations, ability to perform clinical examination (question 4), interpersonal communications (question 7), and availability of the non-teaching staff during the field visits (question 8). More than 80% of students felt that community exposure through this program will assist them to improve their communication skills, understand the various socio-demographic factors associated with the common diseases, apply the same knowledge in treatment, and will enable them to respect the local culture during their practice (questions 12-15)

**Figure 4:**
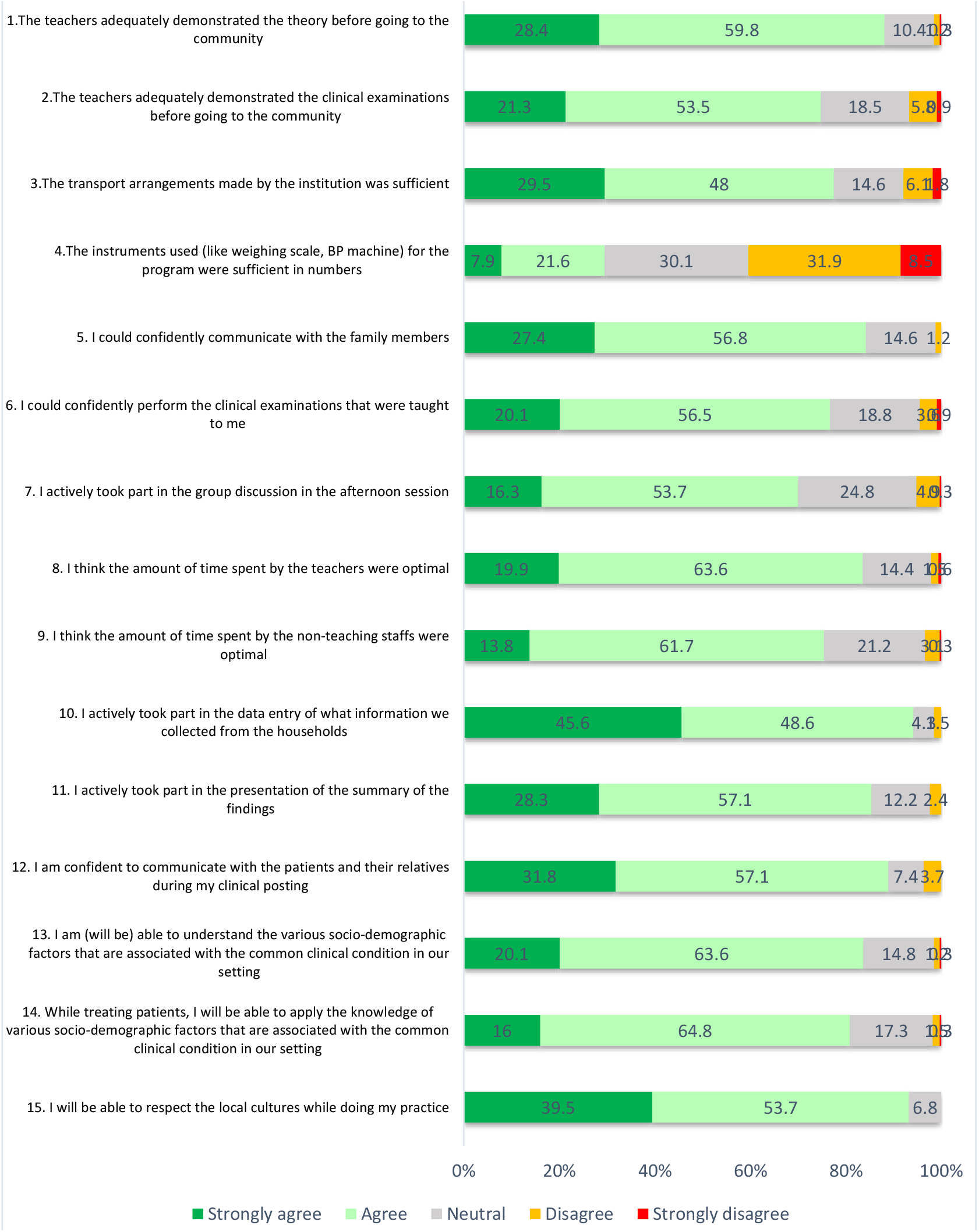
Feedback by the students (n=332)

## Discussion

In this paper, the methods of adopting a relatively new mode of community-based medical education in an Indian setting were presented. The systematic approach on how the undergraduate medical students were exposed to some basic learning experiences within their core subject in a real-life situation, as guided by the guidelines laid down by the apex educational body in the country were described. The COP has provided a unique platform for the students to learn based on several complementary learning theories.

The introduction of the students to the community helped to get to know one another, which proofed to be a key first step in the bonding process between the two parties. It supported their understanding of the social factors, like norms, cultures, and behaviours of patients which influence medical practice. This was strengthened by an awareness of other social factors like recreation, income and expenditure, hygiene, micro- and macro environment, child rearing, pregnancy, and various disease conditions. The knowledge gained by the students were further consolidated during-small group teaching, and during the final presentations. A key benefit of the program was the students’ motivation to learn more about these factors during future learning. For example, one pair observed a physically and mentally challenged child within the community. They learned from the family that the child had a rare genetic condition, as informed by their doctor, and the child often suffers with infections. Due to economic constraint, the family faced challenges to meet the medical and social needs of the child. The students also noticed the emotional state of the parents during the interview. In the group discussion, the pair shared their experience with their fellow students. The group learned from the teachers that there are national programs in existence which addresses such conditions. Subsequently, during the grand presentation, other interesting cases were presented as well. Following the presentations, the village administrators and the local health workers were notified to provide assisting for needy families identified during the program. Based on their experience gained on the program, it is expected that, in future, when students face a health condition, that they will be able to understand the social background of the conditions, distress of the family, examine and treat the patient compassionately, and will be able to guide the family appropriately for further assistance. In an Indian setting, the doctor-patient relationship is often experienced as difficult, (20,21) and such an educational approach may help in the longer term to reduce the doctor-patient factors which deteriorate the relationship.

Importantly, as students had limited subject knowledge, it would have been challenging to teach them new medical knowledges. Therefore, the topics were chosen in such a way that only a minor understanding of the medical field was required, and focus on their understanding of community factors which influence the practice of medicine. It was also anticipated that, due to the different socio-economic backgrounds of the students, exposing the students to the community would assist in them bridging the fear of working in a new environment, and also create the opportunity to learn from each other as students, and to motivate them to learn new theory. For example, in the class-room situation they were taught that ideally, new-born babies should be given only breast milk up to the age of six months (exclusive breastfeeding). When a pair was informed in the community that a new-born baby was given honey and water as pre-lacteal feed instead of breast milk, they shared the experience in the small group session. Another student shared that this is a common practice in her native place as well. With a quality discussion amongst the students and the teacher, the students learned the importance and benefits of breastfeeding, and the common myths in different communities relating to it. They were also informed about the harmful effects of pre-lacteal feeding. These collaborative learning experiences are not possible in classroom teaching at this early stage of their professional education.

Similarly, for the development of clinical skills, the students could only be taught a few clinical examination techniques in the classroom setting. The benefit of the clinical examinations was three-fold. Firstly, the students gained confidence in examining a patient. Secondly, they understood the importance of interpreting their findings. For Example, a pair found that the blood pressure of a known hypertensive person was within normal range. They raised a question based on their classroom learning that blood pressure should be high among hypertensive patients. The teacher then explained the role of medicine and a healthy lifestyle in controlling blood pressure. Lastly, it can enhance their motivation to learn more clinical skills in future.

The importance of data was also introduced to the students. Apart from understanding the individual or family-level information, they experienced the importance of collective data during data entry, analysis, and how these are clustered under different themes. With simple statistics, they came to know the collective information about the community. The collation of data from the different groups and presenting it under various themes helped the students in learning overall findings which they may not have understood if they would have been exposed only to their own observations.

Despite the successes described above, some challenges were also experienced. Apart from administrative challenges insufficient logistics and instruments described before, there were academic hurdles embedded within the teaching-learning process. With the growing change in teaching and learning methods, teachers needs to be continuously trained and retrained to provide quality teaching. Although this is supported by the MCI, who provides training to the medical teachers, (22) the adequacy of such training for the teachers is yet to be evaluated. The goals of these educational achievements may also get diminished over time unless they are appropriately designed, as well as horizontally integrated into the educational approach by involving the various departments during subsequent postings of the UGs in the Community Medicine department. Additionally, the uniformity in curriculum also lacked because of the variations in the rules and regulations amongst the different universities. (11) However, with the introduction of Graduate Medical Education Regulations in 2018, it is expected that these diversified approaches will be addressed in the undergraduate programs. Lastly, the effect of the recent COVID-19 pandemic also put these community-based educational activities at risk. (23) An online mode of teaching cannot offer the various learning experiences as provided by such a program.

## Conclusion

Early initiation of CBME in the form of community orientation program in an Indian setting shows promising results, as it offers multiple advantages in facilitating the teaching process envisioned by the MCI. The COP is expected to touch upon most of the national- and institutional-level objectives envisioned by the Medical Council of India for the Indian medical graduates (IMG). Primarily, students will be able to conceptualize the “health for all” concept and the health rights of all the citizens within this framework of the teaching-learning process. As a part of the foundation course, and within a short time, the holistic approach of the COP showed a potential to facilitate the further learning process of the Indian Medical Graduates by imparting knowledge, attitude, skills, values, responsiveness, and the concept of ethics; and proposes to develop a sensitive and patient-orientated doctor in future. But importantly, all stakeholders – academic and non-academic -should collectively find solutions to overcome the various challenges and foster such programs to achieve its goals.

## Data Availability

We cannot provide the data

